# Platelet Dependent Thrombogenicity, Inflammation and Five Year Mortality in Patients with Non-ST Elevation Acute Coronary Syndrome and Type 2 Diabetes Mellitus: An Observational Study

**DOI:** 10.1101/2022.03.04.22271893

**Authors:** Girish N Viswanathan, Andrew R. Harper, Juan J Badimon, Sally M Marshall, Azfar G Zaman

## Abstract

Platelet dependent thrombogenicity is higher in type 2 diabetes mellitus (T2DM) but there is no data on association between thrombus and mortality or vascular inflammation.

We studied 80 patients who received guideline based therapy (40 T2DM) after NSTE-ACS. Platelet dependent thrombus was assessed using the validated ex vivo Badimon Chamber and all patients were followed up for five years. Bio-markers of coagulation and inflammation were measured.

There were 17 (21.3%) deaths in total (12 in the T2DM, 71% of all deaths), at 5 years. Of the patients who died, thrombus burden was higher (µ^2^ per mm, Mean ± SD, 12,164 ± 3235 vs 9,751 ± 4333). Serum inflammatory cytokines, P selectin and soluble CD40 ligand were higher in T2DM. Univariate analysis showed thrombus area, diabetic status, age, interleukin 1 and P selectin were independent predictors of mortality. Inflammatory biomarkers were associated with thrombus burden. If these findings can be proven in large scale studies, this will lead on to novel therapeutic strategies especially in the current Covid 19 pandemic which has renewed our interest of this interplay of pathophysiology between thrombus and inflammation.

## Introduction

Thrombus formation over a ruptured atheromatous plaque is central to the pathophysiology of acute coronary syndromes. Vascular inflammation plays a key role in plaque events in high risk individuals. Whole blood platelet dependent thrombogenicity using ex vivo clotting chambers has demonstrated differences in patients with and without diabetes [1,2]. However, no study has shown a clear association between thrombus burden and subsequent all-cause mortality or inflammation. Covid 19 pandemic has exposed the interplay of inflammation and thrombosis in these high-risk individuals and there is a renewed interest to understand the pathology in more detail [3,4].

Our primary objective was to explore the association between thrombus quantity and subsequent mortality in T2DM patients after non ST elevation acute coronary syndrome. The secondary aim is to study the role of inflammation bio markers in these individuals.

## Methods

Patient selection: Patient recruitment details and study methodology have been published elsewhere [1]. Forty patients with T2DM and 40 without T2DM were studied one week after NSTE-ACS. The study was approved by Sunderland Research Ethics committee and all patients gave written informed consent. The study is registered with www.clinicaltrials.gov (NCT00728286) and adopted as a portfolio study by the National Institute of Healthcare Research, United Kingdom (UKCRN 7338, www.ukcrn.org).

### Eligibility criteria

All patients had clinical features suggestive of cardiac ischaemia and elevated high sensitive cardiac troponin I (>0.1 μmol/L) on at least 2 occasions 12 hours apart. Furthermore, all patients underwent coronary angiography and only those with documented coronary artery disease with at least one major vessel with >50% stenosis and had percutaneous coronary revascularisation were included.

Patients with T2DM were defined as either a) taking glucose lowering medications, b) with HbA_1c_ >6.5% and or random blood glucose >11.1 mmol/l or d) with fasting blood glucose >7.0 mmol/l, measured on two occasions (All patients were treated as per AHA/ESC guidelines on management of NSTE-ACS^18^ and received aspirin (300 mg loading and 75 mg daily maintenance) and clopidogrel (300 mg loading and 75 mg daily maintenance). In addition, patients received all three standard secondary prevention medications: hydoxymethyl co-enzyme A reductase inhibitor (HMG-CoA), angiotensin converting enzyme (ACE) inhibitor (or angiotensin receptor blocker when clinically indicated) and a beta-blocker (or a calcium channel blocker if clinically indicated). All patients with T2DM were reviewed by the local diabetes specialist team and were treated with oral agents and or insulin as appropriate.

Exclusion criteria were patients with angiographically normal coronary arteries, patients who had coronary artery bypass surgery, current smokers, those receiving glycoprotein IIb/IIIa inhibitor therapy during admission or treated with antiplatelet drugs other than aspirin and clopidogrel. Patients on warfarin, with pre-existing coagulation abnormalities or anaemia (Hb<12g/dL) and known to have malignancy or being investigated for malignancy, were also excluded. The study was performed in a tertiary centre hospital with patients referred from 8 secondary care centres.

Mortality and outcomes data were collected from clinic visits and the UK Office of National Statistics (ONS, www.ONS.org) for 5 years till 2018. The data from ONS is independently verified and this collects all cause mortality. All deaths were attributed to coronary artery disease, but it is unknown if the primary cause of death was solely cardiac in origin.

### Thrombus assessment

Patients received once daily clopidogrel 75 mg and aspirin 75 mg following an initial 300 mg loading dose of both agents. Description of patient selection and thrombus assessment are published elsewhere [1]. Blood thrombogenicity was assessed within 6-10 days after NSTE-ACS using the high shear ex-vivo perfusion (Badimon) chamber as previously described. Briefly, the high shear chambers (inner lumen diameter of 0.1mm, Reynolds number 1690 s^-1^) simulate flow conditions in a moderately stenosed coronary artery. We used surgically dissected porcine aorta as a surrogate for deep arterial injury following plaque rupture. The chamber was maintained in a water bath at 37°C. Native (non-anticoagulated) blood was allowed to flow from an antecubital vein into the perfusion chamber regulated by a distal peristaltic pump at a rate of 10mls min^-1^, using 14.0 Tygon® tube. (Cole Palmer, IL, USA).

### Biomarkers assessment

Serum inflammatory markers, tumour necrosis factor α (TNFα), interleukin 6 (IL-6), high sensitivity C reactive protein (hsCRP), interleukin 1 (IL-1) and interferon gamma (IFNγ) were measured by electro-chemoluminescence (ECL) method (Meso Scale Devices, Gaithersburg, MD, USA) [5,6]. Platelet activation markers serum P–selectin and soluble CD40 ligand levels were measured in serum using sandwich ELISA methods (R&D systems, Abington, UK).

Thrombus assessment and serum inflammatory markers were performed at baseline but not repeated at 5 years as serial assessment of these values were beyond the scope of the study.

Statistical analysis was performed on SPSS v17.1 (SPSS Inc, NY, USA). Data were expressed in mean and median as appropriate. Student t test and Mann Whitney U test were used where appropriate with Spearman rho test used for correlation statistics. For multiple group analysis, one-way ANOVA with post hoc t-test, Bonferroni’s correction was applied.

This observational, single centre study was conducted in accordance with principles of Declaration of Helsinki. All patients gave written informed consent prior to all study specific procedures.

## Results

Baseline demographics were published before [1] as shown in Appendix Table A and baseline data on inflammation and platelet activation biomarkers are shown in Table 1.

**Table A:** Baseline Characteristics of participants with and without type 2 diabetes mellitus.

|  | T2DM (n=40) | Non T2DM (n=40) | P value |
| --- | --- | --- | --- |
| **Demographic data Mean ± SD or %(n)** |  |  |  |
| Age, years | 63.8 ± 12.2 | 62.2 ± 11.4 | 0.553 |
| Male gender, % (n) | 67.5 (27) | 90.0 (36) | 0.027 |
| Body mass index, kg/m<sup>2</sup> | 33 ± 8.3 | 30 ± 5.0 | 0.029 |
| Waist to hip ratio | 1.0 ± 0.05 | 1.0 ± 0.05 | 0.422 |
| Systolic BP, mmHg | 130 ± 19 | 125 ± 20 | 0.212 |
| Diastolic BP, mm Hg | 73 ± 12 | 69 ± 8 | 0.137 |
| **Risk profile: %(n)** |  |  |  |
| History of hypertension | 77.5 (31) | 45.0 (18) | 0.003 |
| History of dyslipidaemia | 90.0 (36) | 42.5 (17) | 0.001 |
| History of chronic kidney disease | 27.5 (11) | 15.0 (6) | 0.172 |
| History of acute myocardial infarction | 27.5 (11) | 12.5 (5) | 0.094 |
| **Medications: %(n)** |  |  |  |
| B blockers | 82.5 (33) | 97.5 (39) | 0.057 |
| Calcium channel blockers | 37.5 (15) | 17.5 (7) | 0.055 |
| **Laboratory data: mean ± SD** |  |  |  |
| HbA1c, % | 7.5 ± 1.5 | 5.8 ± 1.4 | |
| Random plasma glucose, mmol/l | 11.2 ± 6.8 | 6.3 ± 1.7 | |
| Fasting plasma glucose, mmol/l | 7.8 ± 3.1 | 5.2 ± 0.8 | |
| Haemoglobin, g/dl | 13.2 ± 1.8 | 14.0 ± 1.3 | 0.027 |
| Platelets, x1000/mm<sup>3</sup> | 254 ± 89.9 | 246 ± 68.2 | 0.685 |
| Fibrinogen, g/l | 4.2 ± 0.8 | 3.5 ± 1.4 | 0.012 |
| S Creatinine, μmol/l | 111 ± 65.0 | 104 ± 28.3 | 0.560 |
| HsCRP, mg/l | 10.9 ± 9.7 | 3.6 ± 3.2 | 0.001 |
| Total cholesterol, mmol/l | 3.4 ± 1.0 | 3.8 ± 0.9 | 0.135 |
| LDLc, mmol/l | 1.6 ± 0.6 | 1.9 ± 0.9 | 0.023 |
| HDLc, mmol/l | 1.0 ± 0.3 | 1.1 ± 0.4 | 0.301 |
| Triglyceride, mmol/l | 1.9 ± 1.2 | 1.5 ± 0.6 | 0.048 |
| Troponin I, μg/l | 6.9 ± 10.2 | 7.0 ± 12.7 | 0.974 |
| **Note:** Differences in baseline characteristics between patients with and without type 2 diabetes mellitus (T2DM) are shown in the table. HsCRP: High sensitivity C Reactive Protein, LDL: Low Density Lipoprotein, HDL: High Density Lipoprotein. |  |  |  |

**Table 1:** Baseline biomarkers of platelet activation and inflammation of participants with and without type 2 diabetes mellitus (T2DM)

|  | T2DM (n=40) |  | Non T2DM (n=40) |  |  |
| --- | --- | --- | --- | --- | --- |
| | Mean ± SD | Median (IQR) | Mean ± SD | Median (IQR) | P value |
| Fibrinogen, g/l | 4.2 ± 0.8 | 4 (4-5) | 3.5 ± 1.4 | 3 (1-5) | 0.012 |
| P selectin, ng/ml | 76.1 ± 25.6 | 76 (57-93) | 56.7 ± 20.5 | 52 (42-69) | 0.001 |
| Soluble CD40 ligand, ng/ml | 5991 ± 5258 | 4208 (2042-7186) | 2268 ± 2960 | 1233 (593-2839) | 0.001 |
| PAI-1, ng/ml | 96.9 ± 45.7 | 93.7 (58.3-136.6) | 69.7 ± 31.3 | 68.2 (43.9-91.8) | 0.016 |
| HsCRP mg/l | 10.9 ± 9.7 | 10.8 (3.1-11.2) | 3.6 ± 3.2 | 2.8 (2.0-3.9) | 0.001 |
| IFNγ ng/ml | 7.4 ± 10.8 | 4.7 (3.2-7.9) | 4.6 ± 3.8 | 3.5 (2.9-4.7) | 0.131 |
| IL-1 ng/ml | 3.1 ± 13.0 | 0.4 (0.1-2.2) | 0.9 ± 0.6 | 0.8 (0.5-1.3) | 0.284 |
| IL-6 ng/ml | 3.3 ± 2.4 | 2.2 (1.7-4.9) | 1.8 ± 0.8 | 1.6 (1.5-2.2) | 0.001 |
| TNFα ng/ml | 9.5 ± 3.8 | 9.4 (7.0-10.9) | 7.7 ± 3.3 | 7.9 (4.9-10.0) | 0.024 |

**Note:** Differences in biomarkers and platelet activation markers between patients with and without type 2 diabetes mellitus (T2DM) are shown in the table. Both mean with standard deviation and median with 25-75 interquartile range (IQR) values were shown as both normal and non-normal distributed parameters were included in the table.
PAI: Platelet Activation Inhibitor; HsCRP: High sensitivity C Reactive Protein, IFNγ : Interferon gamma, IL: Interleukin, TNFα : Tumour Necrosis Factor alpha.

At five years follow up, there were seventeen deaths overall (21.3%) of which 12 were T2DM and 5 were non DM (71% vs. 29% of all deaths, p=0.056). In patients who died, thrombus burden was higher (µ^2^ per mm, Mean ± SD, 12,164 ± 3235 vs 9751 ± 4333, p=0.036 for low shear and 17756 ± 10583 vs 16675 ± 11720, p=0.716 for high shear).

Patients withT2DM had higher inflammatory and coagulation biomarkers (Table 1) and patients who died had higher baseline inflammatory and coagulation biomarkers (Table 2). When stratified according to presence or absence of T2DM, P selectin, interleukin 6 and TnFα levels were highest in T2DM patients who died (Table 3).

**Table 2:** Mortality and bio-markers.

| Median (25-75 IQR) | Dead (n=17) | Alive (n=63) | P value |
| --- | --- | --- | --- |
| P Selectin ng/ml | 77.7 (56.7-93.3) | 59.4 (45.4-76.1) | 0.031 |
| Soluble CD40 ligand ng/ml | 2817 (1151-7113) | 2239 (1048-5359) | 0.303 |
| Fibrinogen g/l | 3.6 (3.0-4.6) | 4.2 (3.0-4.7) | 0.517 |
| HsCRP mg/l | 5.1 (2.1-11.2) | 3.7 (2.1-10.6) | 0.303 |
| IFNγ ng/ml | 8.3 (6.4-10.2) | 3.8 (3.0-5.5) | 0.221 |
| IL-1 ng/ml | 0.41 (0.06-0.98) | 0.29 (0.19-0.66) | 0.074 |
| IL-6 ng/ml | 1.9 (1.6-4.1) | 1.9 (1.5-2.6) | 0.298 |
| TNFα ng/ml | 9.2 (4.6-12.1) | 8.3 (6.4-10.2) | 0.576 |

**Note:** Differences in biomarkers and platelet activation markers between patients who are alive and deceased are shown in the table.
PAI: Platelet Activation Inhibitor; HsCRP: High sensitivity C Reactive Protein; IFNγ : Interferon gamma; IL: Interleukin; TNFα : Tumour Necrosis Factor alpha.

**Table 3:** Factorial table on mortality and type 2 diabetes mellitus status.

| Mean ± SD | T2DM+dead (n=12) | NonDM+dead (n=5) | T2DM+alive (n=28) | NonDM+alive (n=35) | P value |
| --- | --- | --- | --- | --- | --- |
| Age, in years | 70.4 ± 10.8 | 76.0 ± 11.5 | 61.0 ± 11.8 | 60.3 ± 10.2 | 0.002 |
| BMI | 36.3 ± 9.2 | 30.1 ± 4.1 | 32.5 ± 8.1 | 29.7 ± 5.1 | 0.079 |
| Serum Creatinine μmol/l | 136 ± 13 | 111 ± 22 | 100 ± 22 | 103 ± 29 | 0.189 |
| Total Cholesterol mmol/l | 3.7 ± 1.1 | 3.3 ± 0.8 | 3.3 ± 1.0 | 3.8 ± 0.9 | 0.230 |
| LDLc mmol/l | 1.8 ± 0.8 | 1.4 ± 0.7 | 1.5 ± 0.5 | 2.0 ± 0.7 | 0.025 |
| HDLc mmol/l | 1.2 ± 0.5 | 1.0 ± 0.2 | 1.0 ± 0.2 | 1.1 ± 0.4 | 0.164 |
| Triglycerides mmol/l | 1.6 ± 0.7 | 1.9 ± 1.1 | 2.1 ± 1.4 | 1.4 ± 0.5 | 0.070 |
| Random plasma glucose mmol/l | 15.7 ± 9.6 | 6.5 ± 2.0 | 9.4 ± 4.5 | 6.3 ± 1.7 | 0.001 |
| Fasting plasma glucose mmol/l | 9.3 ± 4.7 | 5.4 ± 0.4 | 7.2 ± 2.1 | 5.2 ± 0.8 | 0.001 |
| HbA1c % | 7.8 ± 1.7 | 5.8 ± 0.3 | 7.4 ± 1.4 | 5.8 ± 1.5 | 0.001 |
| IFNγ ng/l | 7.1 ± 3.7 | 3.6 ± 3.0 | 7.4 ± 12.7 | 4.7 ± 3.9 | 0.503 |
| IL-1 ng/l | 1.9 ± 2.2 | 0.7 ± 0.4 | 4.8 ± 15.2 | 0.9 ± 0.6 | 0.221 |
| IL-6 ng/l | 3.4 ± 2.1 | 1.7 ± 0.7 | 3.3 ± 2.5 | 1.8 ± 0.8 | 0.005 |
| TNFα ng/l | 8.6 ± 4.0 | 9.8 ± 4.1 | 9.9 ± 3.7 | 7.3 ± 3.1 | 0.039 |
| HsCRP mg/l | 10.9 ± 7.0 | 1.9 ± 0.6 | 11.0 ± 10.8 | 3.8 ± 3.4 | 0.001 |
| P selectin pg/l | 84.6 ± 28.4 | 60.8 ± 18.8 | 72.5 ± 23.9 | 56.1 ± 20.9 | 0.002 |
| Soluble CD40 ligand pg/l | 6944.5 ± 5411 | 1128.8 ± 935 | 5582.4 ± 5237 | 2431.1 ± 3120 | 0.002 |
| Fibrinogen g/l | 4.1 ± 1.0 | 2.7 ± 1.1 | 4.3 ± 0.8 | 3.6 ± 1.8 | 0.039 |
| High shear thrombus $\mu^2$/mm | 20667 ± 12213 | 12424 ± 5722 | 19329 ± 10086 | 15104 ± 8872 | 0.100 |
| Low shear thrombus $\mu^2$/mm | 13062 ± 3343 | 10011 ± 1718 | 11276 ± 4722 | 8575 ± 3658 | 0.004 |

**Note:** Differences between patients with and without type 2 diabetes mellitus (T2DM) who are stratified according to alive or dead status are shown in the table. One way ANOVA values are shown with mean and standard deviation.
BMI: Body Mass Index, HsCRP: High sensitivity C Reactive Protein, IFNγ : Interferon gamma, IL: interleukin, TNFα : Tumour Necrosis Factor alpha, LDL: Low Density Lipoprotein, HDL: High Density Lipoprotein.

Those with T2DM who died were 6 years younger at baseline than those without T2DM (mean ± SD, in years, T2DM vs Non DM; 70 ± 10.8 vs 76 ± 11.5, p=0.002). Univariate analysis showed thrombus area (r=0.236, p=0.001), diabetic status (r= 0.366, p=0.050), age (r=0.401, p=0.001), serum creatinine levels (r= 0.222, p=0.050), interleukin 1 (r=0.117, p=0.027) and P selectin (r=0.234, p=0.037) were independent predictors of mortality. However, after adjusting for age, BMI and diabetic status, thrombus area was not an independent predictor of mortality in multivariate analysis.

Inflammatory biomarkers were also associated with thrombus in univariate analysis were serum IL-6 (rho=0.251, p=0.027), serum TNFα (rho=0.319, p=0.005) and serum fibrinogen (rho 0.283, p=0.013). However, these associations were lost on multivariate analysis.

## Discussion

In this study of 80 patients after NSTE-ACS, there was increased 5 year mortality in patients with T2DM and an association of mortality with thrombus quantity. Our study also revealed an association of thrombus with inflammatory cytokines. Due to smaller size of our study population, multivariate analysis could not prove this association to be independent of other risk factors.

Patients with T2DM and NSTE-ACS have higher cardiovascular morbidity and mortality despite current secondary prevention therapy. We have shown previously that whole blood thrombogencity is increased in T2DM after NSTE-ACS [1] and this report assesses its relation to mortality. Several previous reports have demonstrated the utility of the Badimon ex vivo chamber to measure platelet dependent thrombus – in patients with diabetes mellitus [7], after exposure to pollutants [8] and to test for efficacy of antithrombotic medication [9]. In small numbers of patients, the chamber is sensitive in detecting differences in platelet dependent thrombus formation but no study has yet correlated thrombus quantity in the chamber with mortality.

Population studies show ongoing ischaemic cardiac events for several years after ACS [10] and the PEGASUS study showed benefits of longer term dual antiplatelet therapy up to 3 years in high risk individuals [11]. THEMIS study on patients with T2DM and previous history of MI showed reduced cardiovascular events but at the expense of increased bleeding [12]. Persistence of thrombogenicity is likely that a contributory factor in the increased morbidity and mortality, several years after ACS is and our study findings suggest that increased thrombus burden soon after ACS may identify patients susceptible to further ischaemic events beyond the 12 months of recommended dual antiplatelet therapy.

Anti-inflammatory Therapy with Canakinumab for Atherosclerotic Disease (CANTOS) trial [13] has shown for the first time that monoclonal antibody to IL-1β (an upstream driver of the Il-6 pathway) reduced recurrent cardiac ischaemic events after ACS. In light of this, it is reassuring that in our small study we found an association of IL-6 and TNF-α with thrombus. These findings are novel and may imply a simple association secondary to ACS [14] or a causal factor for increased thrombogenicity in these high risk individuals [15].

Persistent inflammation in treated T2DM subjects has been linked to future events and mortality and thus formed the basis for the “common soil” hypothesis [16]. The exact mechanism by which inflammation contributes to thrombogenicity is unknown. Inflammation up-regulates procoagulants, and down regulates anticoagulants and fibrinolysis [17,18].

Indirect evidence shows TNFα promotes thrombogenicity by inducing expression of tissue factor in monocytes, platelet and leucocyte adhesion to endothelium [19], and production of large multimers of van Willebrand factor (vWF). Interleukin-6 induces synthesis of new platelets which are more thrombogenic and contribute to thrombogenicity in T2DM [20]. In addition, high levels of IL-6 delay clearance of vWF thereby sustaining thrombogenicity [21]. Both TNFα and IL-6 also down regulate naturally occurring antithrombotic substances such as protein C and thrombomodulin [22]. Pro thrombotic mediators like P selectin and soluble CD 40 (sCD40L) ligand were higher and provide a possible mechanistic link for high blood thrombogenicity in patients with T2DM [23-26]. Literature on P selectin and sCD40L is minimal in T2DM patients after NSTE-ACS [27].

Findings from our study confirm the persistence of a pro-inflammatory and pro-thrombotic state in T2DM and ACS [28,29] despite current aggressive secondary prevention. Importantly, we demonstrate a direct association to thrombus formation and an indirect association to mortality of these pro-inflammatory and pro-thrombotic markers.

## Conclusions

We have shown that 5-year mortality is higher in patients with T2DM despite standard secondary prevention therapy and mortality is associated with increased whole blood thrombus and inflammatory cytokines. Also, we would like to acknowledge that further studies are required with incorporation of angiographic data and atheroma burden to conclude whether high mortality is due to aggressive atherosclerosis or high thrombogenicity.

## Data Availability

All data produced in the present work are contained in the manuscript

## Contribution of the Authors

G.N. Viswanathan conceived the idea, designed and conducted the study, performed the analysis and interpretation of the data, and drafted the manuscript. A.R. Harper expanded the idea, did analysis and interpretation of the data, drafted the manuscript and provided critical revision. J.J. Badimon interpreted the data, critically reviewed the manuscript. S.M Marshall improved the original idea and design of the study, interpreted the data and critically reviewed the manuscript. A.G. Zaman expanded concept and design of the study, performed analysis of the data, provided quality control for the data, critically reviewed and interpreted the results, and revised the manuscript.

## Acknowledgements

Azfar Zaman was supported by a Clinical Research Fellowship from the British Heart Foundation (BHF), UK (FS/033/07). The BHF and Northumberland, Tyne and Wear Comprehensive Local Research Network funded the study. We acknowledge support from the staff at the Clinical Research Facility, Newcastle upon Tyne Hospital NHS Foundation Trust. We acknowledge the help and support of Dr Karthik Balasubramaniam, Mrs Heather Cook and Ms Annette Lane, Institute of cellular Medicine, Newcastle University for their help in day to day running of the study, biomarker assay and preparation of aorta samples.

